# TClustVID: A Novel Machine Learning Classification Model to Investigate Topics and Sentiment in COVID-19 Tweets

**DOI:** 10.1101/2020.08.04.20167973

**Authors:** Md. Shahriare Satu, Md. Imran Khan, Mufti Mahmud, Shahadat Uddin, Matthew A. Summers, Julian M.W. Quinn, Mohammad Ali Moni

**Author notes:** Corresponding Author Email address (Mohammad Ali Moni).

## Abstract

COVID-19, caused by the SARS-Cov2, varies greatly in its severity but represent serious respiratory symptoms with vascular and other complications, particularly in older adults. The disease can be spread by both symptomatic and asymptomatic infected individuals, and remains uncertainty over key aspects of its infectivity, no effective remedy yet exists and this disease causes severe economic effects globally. For these reasons, COVID-19 is the subject of intense and widespread discussion on social media platforms including Facebook and Twitter. These public forums substantially impact on public opinions in some cases and exacerbate widespread panic and misinformation spread during the crisis. Thus, this work aimed to design an intelligent clustering-based classification and topics extracting model (named TClustVID) that analyze COVID-19-related public tweets to extract significant sentiments with high accuracy. We gathered COVID-19 Twitter datasets from the IEEE Dataport repository and employed a range of data preprocessing methods to clean the raw data, then applied tokenization and produced a word-to-index dictionary. Thereafter, different classifications were employed to Twitter datasets which enabled exploration of the performance of traditional and TclustVID classification methods. TClustVID showed higher performance compared to the traditional classifiers determined by clustering criteria. Finally, we extracted significant topic clusters from TClustVID, split them into positive, neutral and negative clusters and implemented latent dirichlet allocation for extraction of popular COVID-19 topics. This approach identified common prevailing public opinions and concerns related to COVID-19, as well as attitudes to infection prevention strategies held by people from different countries concerning the current pandemic situation.

## 1. Introduction

COVID-19 has become a global concern as a major and dangerous public health threat. The World Health Organization (WHO) declared COVID-19 a Public Health Emergency of International Concern (PHEIC) on February 28, 2020. During the 1960s, coronaviruses (CoVs) were found to infect humans mainly in the upper respiratory tract, most commonly human coronavirus 229E and OC43 [1]. Many CoVs circulate in wild mammalian populations, and cause only minor, if any, human health problems. This picture changed with the emergence of severe acute respiratory syndrome (SARS-CoV) and the Middle East Respiratory Syndrome coronavirus (MERS-CoV) that infect the lung epithelial tissues and cause serious and often deadly respiratory disease [2]. However, SARS-CoV and MERS-CoV outbreaks in 2002 and 2012 respectively receded, probably due to the lack of spread from non-symptomatic individuals that allowed rapid containment. In contrast, SARS-CoV2 which causes pneumonia-like symptoms and cardiovascular complications ranging in severity from undetectable to rapidly lethal. This, coupled with its rapid spread has caused huge economic disruption and personal health fears and uncertainties that have dominated both the news and social media.

The massive use of web and mobile technologies gives opportunities for people to share their opinions about issues affecting them on social media platforms such as Facebook and Twitter. During the COVID-19 pandemic social media has been used both for normal daily interaction and to spread health messages, but there are also significant numbers of messages left by users sharing their general feelings about personal situations, their health status, the care they take to stay well, and much other COVID-19-relevant information [3]. Such messages may provide useful large scale insights into behavioral responses to the pandemic, however it is not easy to judge whether a social media message carries important information, not least because semantic abstruseness makes it hard to understand many messages. Nevertheless, machine learning and computational methods have increasingly been used to scrutinize social media data in the biomedical sector [4]. The content of COVID-19 related messages may be used to extract information that can inform physicians and policy-makers. Twitter, in particular, is a popular microblogging and public networking service widely used for messaging and posting [5]. Automatic classification of tweets into particular classes is challenging, not least because these messages are short, 140 characters, or less [6]. The analysis requires identification of sentiments in Twitter messages (tweets) which contain abbreviations, spelling variations and ambiguous or informal language.

Some recent studies have attempted to scrutinize COVID-19 tweets in bulk for health purposes, although it is likely they have also been mined for commercial purposes. Lopez et al. [7] generated a dataset of multilingual tweets collected from all over the world since January 22^*nd*^. In this dataset they identified common responses and how they changed across time. Kouzy et al. [8] explored tweets using 14 trending hashtags and keywords about COVID-19 and investigated the magnitude of misinformation by comparing terms and hashtags of tweets. Cinelli et al. [9] analyzed the dissemination of information about COVID-19 on Twitter, Instagram, YouTube, Reddit, and Gab, and found a quite different volume of misinformation in each of the platforms. Medford et al. [10] analyzed all twitter user data from January 14^*th*^ to 28^*th*^, 2020 and applied sentiment analysis and topics modeling using LDA to explore discussion topics over time. However, there are few dedicated machine learning based tweet analysis models to investigate user sentiments about COVID-19. In this study, we sourced several twitter datasets and investigated sentiment topics related to COVID-19 by designing a novel clustering based analysis model named TClustVID. This model was used to explore significant subsets (clusters) from COVID-19 twitter datasets and select them by applying the highest classification performance approach. Each of these twitter clusters has been split into the positive, negative and neutral cluster and employed latent dirichlet allocation (LDA) to extract key topics from each of them. Topics were interpreted to identify the most frequent significant topic among the tweets studied. This methodology can be used to generate information relevant to researchers and policymakers when dealing with COVID-19 issues that relate to the general public and human social behaviour at large.

## 2. Materials and Methods

We proposed a machine learning based COVID-19 tweets analytic model that can be used to explore significant topics from Twitter datasets. To process different types of tweets, several natural language processing techniques are used, along with machine learning methods as illustrated in Figure 1.

**Figure 1:**
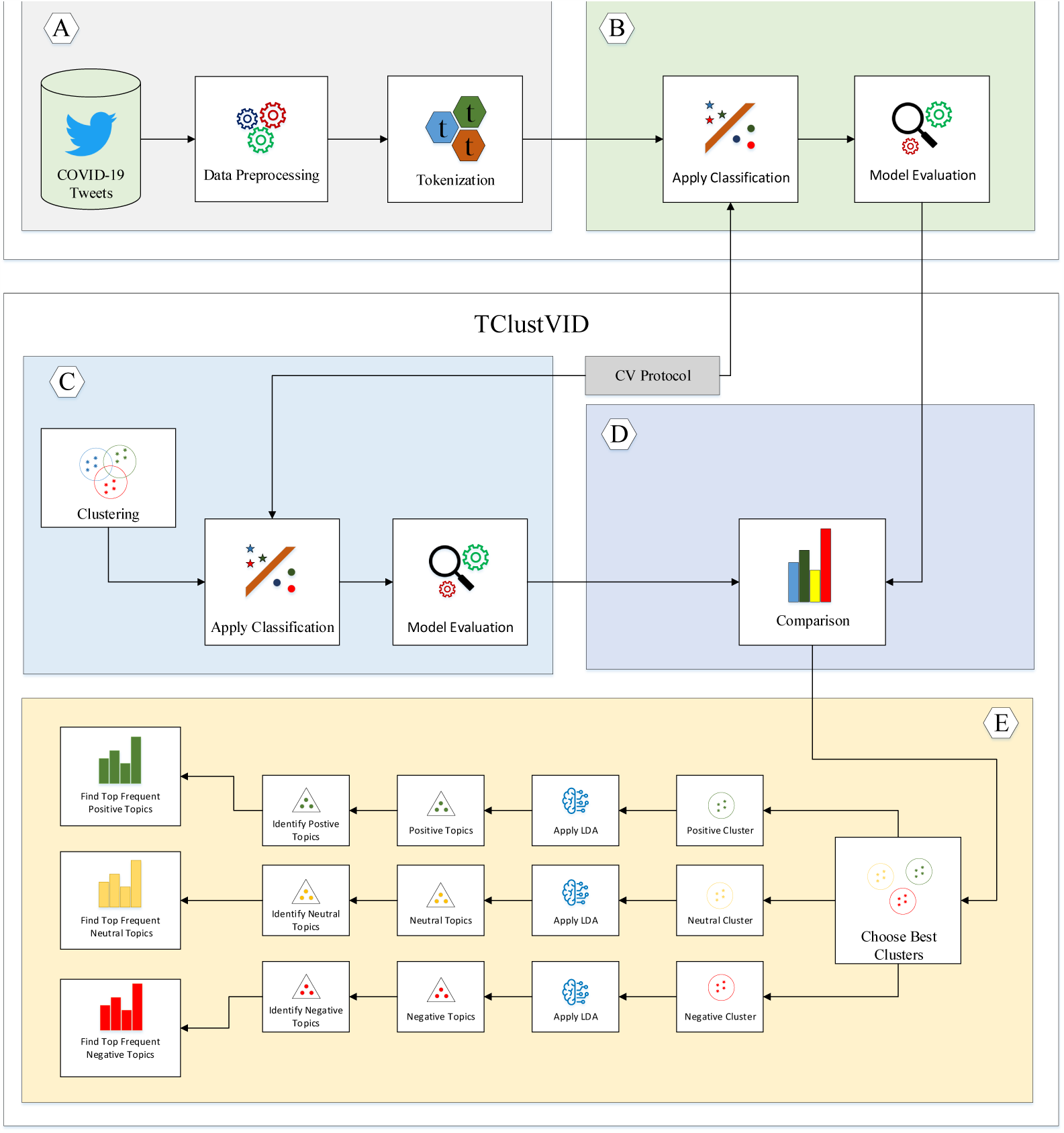
Details of Working Methodology where A. Data preprocessing B. Traditional classification and evaluation C. Clustering, classification and c valuation D. Comparison the outcomes between traditional and TClustVID and Select the best clusters E. Identify positive, neutral and negative clusters, extract topics by LDA and represent top frequent topics from it

### 2.1 Data Description

COVID-19 twitter datasets were collected from the IEEE Data portal that originated from the LSTM model, developed by Rabindra Lamsal, which monitors the real-time twitter feed for COVID-19-related tweets [11]. It generates over 0.3 million requests every 24 hours and its time-series graph is updated every 30 seconds. Almost 16 million tweets were identified before March 20^*th*^ 2020. Each database (*.db) contains three attributes in which first, second, and the third columns denote the date and time of the tweet, and the tweets and sentiment scores. However, these sentiment scores are manipulated within the range [0,2] where the most negative, neutral, and positive sentiment are indicated as 0, 1 and 2, respectively. Eight twitter datasets (corona tweets 1M.db, corona tweets 1M 2, corona tweets 1M, corona tweets 2L, corona tweets 2M.db, corona tweets 2M 2, corona tweets 2M 3 and corona tweets 3M) have been investigated and deemed suitable models to classify tweets in this study. Each dataset has been denoted the tweets related to COVID-19 of each day before March 20^*th*^ 2020. We gathered datasets of a couple days to understand and extract various topics everyday. The first seven of these datasets are denoted as dataset-1, dataset-2, dataset-3, dataset-4, dataset-5, dataset-6, and dataset-7. In this study, corona tweets 3M was split into dataset-8 and dataset-9 because the computational cost is manipulated very high for the corona tweets 3M.

### 2.2 Data Preprocessing

In preprocessing steps, different twitter datasets have been prepared to manipulate them. Tweets contain various HTML tags, punctuation, numbers, single characters and multiple spaces. Several functions were used to clean datasets in this step. The symbols ‘ <>′ were replaced with empty spaces. Each of the single characters was replaced with a space as the single letters do not indicate any meaningful communication. Finally, all multiple spaces were removed from these tweets. This process was employed in the nine twitter datasets and combined for further analysis. Table 1 represents the number of tweets before and after prepossessing steps.

**Table 1:** Number of Cleaned Tweets COVID-19 After Data Preprocessing

| Primary Dataset | # tweets (N=19797541) | Denoted | # tweets (N=19712979) |
| --- | --- | --- | --- |
| Before Preprocessing |  | After Preprocessing |  |
| corona_tweets_1M.db | 1578957 | Dataset-1 | 1569619 |
| corona_tweets_1M_2 | 1889781 | Dataset-2 | 1880297 |
| corona_tweets_1M | 1903768 | Dataset-3 | 1894526 |
| corona_tweets_2L | 280304 | Dataset-4 | 276566 |
| corona_tweets_2M.db | 2322153 | Dataset-5 | 2312104 |
| corona_tweets_2M_2 | 2268634 | Dataset-6 | 2257529 |
| corona_tweets_2M_3 | 2081576 | Dataset-7 | 2072575 |
| corona_tweets_3M | 7472368 | Dataset-8 | 3724882 |
|  |  | Dataset-9 | 3724881 |

### 2.3 Tokenization

After preprocessing steps, tokenization procedures were used to generate a word-to-index dictionary whereby each word is created as a key in the corpus. Hence, the corresponding unique index indicates the value of the keys. In the training phase, each list holds each sentence where the size is dissimilar. Thus, the maximum length of each list is fixed. If the length of any list is exceeded, it is truncated to the maximum permitted length. Zeroes are added to the endpoint of a shortlist until it reaches maximum length, a process called padding. Thus, Glove embedding tokenization [12] has been used to create a dictionary that holds a word as a key and the corresponding list as values. Finally, an embedding matrix is generated whereby each row number matches the index of the word in the corpus. Raw tweets contain text instances which cannot handle by machine learning procedure. Therefore, we run data pre-processing and tokenization process to make it executable for clustering and classification computation.

### 2.4 Traditional Approach

After manipulating data preprocessing and tokenization, we implemented different machine learning baseline classifiers into twitter datasets and evaluate the results. This process is called tradition approach. It is a general process to apply classifiers into the dataset. In this work, we implement various well known classifiers into traditional way in the dataset and compare the results with TClustVID. This procedure is used to justify the performance of TClustVID and assist to explore best clusters comparing other methods. However, both traditional and TClustVID use same baseline classifier which are indicated at section 2.6.

### 2.5 TClustVID: Clustered Based Classification and Topics Modeling Approach

We proposed a novel clustering-based topics modeling approach called TClustVID which represents at 1. It splits the twitter datasets into several clusters (groups) applying k-means clustering algorithms. It is implemented into COVID-19 twitter datasets following preprocessing and tokenization process. Clustering is an unsupervised method to find homogeneous groups from the dataset. This procedure is used to create clusters as features and improve classification results. There are remaining various clustering algorithms such as k-means, k-medoids, fuzzy C-means, hierarchical clustering, and density based clustering [13, 14]. K-medoids is not the best choice for analyzing sparse data like tweets. Besides, fuzzy C-means is useful to the sheer volumes of tweets and contains low scalability where human annotation really expensive. The performance of hierarchical clustering is slower than k-means. Density based clustering is highly efficient for clustering unstructured data and less prone to outliers and noise. In this work, we handle a large amount of tweet data where K-means defines the mean point within the cluster by optimizing the Euclidean distance between each instance and cluster mean in a less time [15, 14]. The default values of k are taken as 5 which is also used more frequently this type of work. Each cluster contain positive, negative and neutral tweets. When the clusters are found, the tokens were replaced by primary tweets and re-tokenized each cluster. Then, baseline classifiers were used to investigate the performance of different datasets and extracted clusters using 10 fold cross-validation. Different evaluation metrics such as accuracy, area under the curve (AUC), f-measure, g-mean, sensitivity and specificity have been used to investigate these results.

#### Algorithm 1

TClustVID: Clustered Based Proposed Classification and Topics Modeling Approach

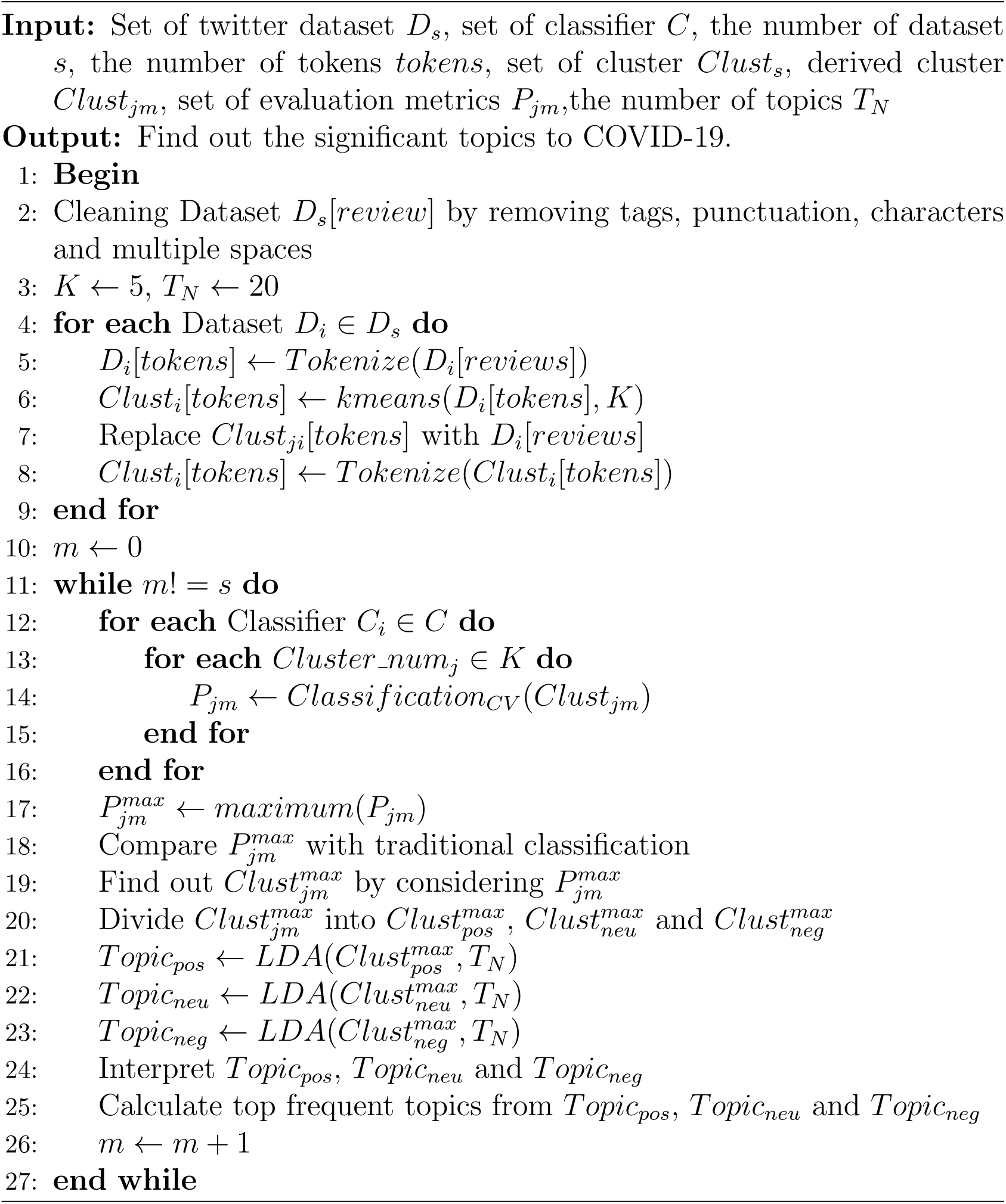

Compared to the classification results of traditional approach and TClustVID, the best performing clusters represent more frequent topics because they show the highest classification performance relative to the traditional approach. These clusters are divided into positive, neutral and negative clusters for further analysis. Therefore, LDA was used to explore significant topics of positive, neutral and negative clusters from the high performing nine clusters. There were extracted 20 topics from each cluster. We represent individual topics into word cloud where each topics contain different words/tokens. In addition, each word cloud represent individual words into different sizes because they organize words according to the weights of them.

But, LDA cannot interpret these topics, hence, we manually analyze the words/tokens of each topics and interpret them.

### 2.6 Baseline Classification

In previous studies, various classifiers such as decision tree (DT), Gradient Boosting (GB), K-Nearest Neighbor (KNN), Logistic Regression (LR), Multi-Layer Perceptron (MLP), Naïve Bayes (NB), Random Forest (RF), Support Vector Machine (SVM) and XGBoost (XGB) have been commonly used to investigate different types of tweet datasets for sentiment analysis. These classifiers were used in similar kinds of twitter data analysis such as C5.0 (DT), KNN, SVM, LR and ZeroR [16], personality prediction using KNN, NB, SVM, and XGB [17, 18], spam detection using RF, NB, SMO and Ibk (KNN equivalent) [19], sentiment analysis using NB, SVM, and MLP of top colleges [20], prediction of alternation price fluctuation using GB [21]. Following this tasks, we selected them to classify the COVID-19 twitter dataset, then explored the best clusters using 10-fold cross-validation.

### 2.7 Evaluation Metrics

A confusion matrix is specified for the performance of the classifier that indicates the number of correct and incorrect predictions when considering known true values. Based on positive and negative classes, it denotes True Positive (TP), True Negative (TN), False Positive (FP) and False Negative (FN).

- Accuracy: It represents the efficiency of the algorithm in terms of predicting true values that is shown in the following equation.

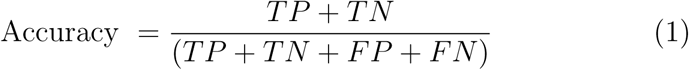
- AUC: It is used to explore machine learning models considering the TP and TN rates represent how well positive classes are isolated from negative classes.

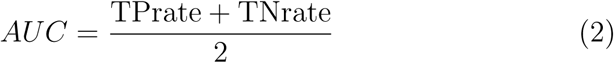
- F-measure: It represents the harmonic mean of the precision and recall which shows the following equation.

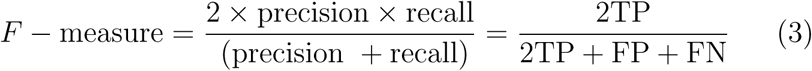
- Geometric-mean (G-mean): It specifies the root of the class-specific sensitivity product and makes a trade-off between the expansion of accuracy on each class and balancing accuracy.

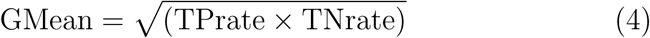
- Sensitivity: The portion of appropriately detected actual positives is indicated as sensitivity using the following equation.

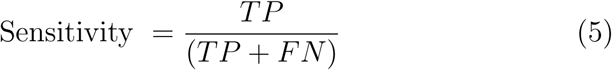
- Specificity: The portion of correctly identified actual negatives is denoted as specificity which represents by the following equation.

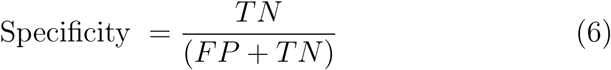

## 3. Experimental Result & Discussion

### 3.1 Classification Approach

Various classification algorithms were used to analyze the COVID-19 twitter dataset, using the sci-kit-learn machine learning python library [22]. In this study, we have proposed a clustering based classification and topics extraction model, TClustVID, which detects positive, negative, and neutral tweets more accurately than previous methods which allows to explore more significant thematic topics. COVID-19 twitter datasets were cleaned using the data preprocessing procedures described above. Word-to-index dictionaries were then created using GloVe embedding tokenization. Several classification algorithms such as DT, GB, KNN, LR, MLP, NB, RF, SVM and XGB were analyzed sentiments of the COVID-19 datasets, using 10 fold cross validation approach. The experimental analyses of COVID-19 twitter datasets (from dataset 1 to 9) are represented at Table 2 to 10. We used various evaluation metrics such as accuracy, area under the curve (AUC), f-measure, g-mean, sensitivity and specificity to profile the results of the nine COVID-19 twitter datasets used with this model.

**Table 2:** Experimental Result of Dataset-1

| Classifier | Traditional Approach |  |  |  |  | TClustVID |  |  |  |  |
| --- | --- | --- | --- | --- | --- | --- | --- | --- | --- | --- |
|  | Accuracy | AUC | F-Measure | Sensitivity | Specificity | Accuracy | AUC | F-Measure | Sensitivity | Specificity |
| DT | 0.9154 | 0.9009 | 0.9155 | 0.9154 | 0.8864 | 0.9518 | 0.9449 | 0.9518 | 0.9518 | 0.9381 |
| GB | 0.7876 | 0.6526 | 0.7464 | 0.7876 | 0.5176 | 0.8162 | 0.7133 | 0.7902 | 0.8162 | 0.6104 |
| KNN | 0.9098 | 0.8801 | 0.9085 | 0.9098 | 0.8504 | 0.9456 | 0.9298 | 0.9451 | 0.9456 | 0.9141 |
| LR | 0.6954 | 0.5016 | 0.5759 | 0.6954 | 0.3078 | 0.6786 | 0.5004 | 0.5516 | 0.6786 | 0.3222 |
| MLP | 0.8403 | 0.7660 | 0.8303 | 0.8403 | 0.6918 | 0.9014 | 0.8686 | 0.8993 | 0.9014 | 0.8357 |
| NB | 0.6537 | 0.5030 | 0.5965 | 0.6537 | 0.3524 | 0.6438 | 0.5015 | 0.5769 | 0.6438 | 0.3592 |
| RF | 0.9235 | 0.8982 | 0.9226 | 0.9235 | 0.8728 | 0.9573 | 0.9444 | 0.9570 | 0.9573 | 0.9315 |
| SVM | 0.7568 | 0.5996 | 0.6939 | 0.7568 | 0.4424 | 0.8029 | 0.6928 | 0.7718 | 0.8029 | 0.5827 |
| XGB | 0.7871 | 0.6573 | 0.7496 | 0.7871 | 0.5274 | 0.8535 | 0.7742 | 0.8399 | 0.8535 | 0.6949 |

In traditional approaches (see Table-2, 3, 4, 5, 6,7, 8, 9 and 10), RF gave, respectively, 3, 7, 5, 7, 3 and 7 times the highest accuracy, AUC, f-measure, sensitivity, and specificity, respectively, in different twitter datasets. Alternatively, DT gave 6, 2, 2, 2, 6 and 2 times the highest accuracy, AUC, f-measure, sensitivity, and specificity respectively. If the frequency of generating the best values are calculated, RF showed a total 32 times higher results to analyze twitter datasets. However, DT provided total of 20 times the highest results corresponding to RF. It was also noted that DT was better at predicting true positive instances compared to RF. Both of these showed 90% average results to scrutinize COVID-19 twitter datasets. Without DF and RF, KNN and MLP showed better results than other classifiers from dataset-1 to 9. However, the performance of KNN was better than MLP at all times and MLP showed better results than XGB, GB, LR and SVM. Thus, they were considered as the third and fourth top-performing classifiers correspondingly. In the comparison of GB and XGB, most of the time XGB showed better results than GB. XGB gave 5 times higher results when compared to GB. In some cases, GB showed a better result than XGB for only a few metrics (e.g., better accuracy, specificity in dataset-1, better accuracy and sensitivity in dataset-8 and 9). Hence, XGB was the 5^*th*^ high performing classifier in this work. SVM and LR did not demonstrate sound outcomes in analyzing the COVID-19 datasets. For most of these cases, LR gave higher results than SVM.

**Table 3:** Experimental Result of Dataset-2

| Classifier | Traditional Approach |  |  |  |  | TClustVID |  |  |  |  |
| --- | --- | --- | --- | --- | --- | --- | --- | --- | --- | --- |
|  | Accuracy | AUC | F-Measure | Sensitivity | Specificity | Accuracy | AUC | F-Measure | Sensitivity | Specificity |
| DT | 0.9309 | 0.8985 | 0.9310 | 0.9309 | 0.8661 | 0.9637 | 0.9425 | 0.9637 | 0.9637 | 0.9212 |
| GB | 0.8158 | 0.5669 | 0.7550 | 0.8158 | 0.3180 | 0.8563 | 0.6260 | 0.8192 | 0.8563 | 0.3956 |
| KNN | 0.9238 | 0.8651 | 0.9220 | 0.9238 | 0.8064 | 0.9575 | 0.9179 | 0.9569 | 0.9575 | 0.8783 |
| LR | 0.7866 | 0.5014 | 0.6953 | 0.7866 | 0.2162 | 0.8072 | 0.5052 | 0.7269 | 0.8072 | 0.2032 |
| MLP | 0.8673 | 0.7297 | 0.8543 | 0.8673 | 0.5922 | 0.9245 | 0.8405 | 0.9212 | 0.9245 | 0.7564 |
| NB | 0.2132 | 0.5002 | 0.0758 | 0.2132 | 0.7871 | 0.1924 | 0.5002 | 0.0626 | 0.1924 | 0.8079 |
| RF | 0.9374 | 0.8877 | 0.9361 | 0.9374 | 0.8380 | 0.9676 | 0.9362 | 0.9672 | 0.9676 | 0.9048 |
| SVM | 0.7867 | 0.5009 | 0.6947 | 0.7867 | 0.2151 | 0.8063 | 0.5018 | 0.7238 | 0.8063 | 0.1972 |
| XGB | 0.8196 | 0.5775 | 0.7635 | 0.8196 | 0.3355 | 0.8745 | 0.6936 | 0.8547 | 0.8745 | 0.5128 |

**Table 4:** Experimental Result of Dataset-3

| Classifier | Traditional Approach |  |  |  |  | TClustVID |  |  |  |  |
| --- | --- | --- | --- | --- | --- | --- | --- | --- | --- | --- |
|  | Accuracy | AUC | F-Measure | Sensitivity | Specificity | Accuracy | AUC | F-Measure | Sensitivity | Specificity |
| DT | 0.9107 | 0.9304 | 0.9107 | 0.9107 | 0.9501 | 0.9602 | 0.9666 | 0.9601 | 0.9602 | 0.9730 |
| GB | 0.6986 | 0.7168 | 0.6744 | 0.6986 | 0.7351 | 0.8463 | 0.8380 | 0.8360 | 0.8463 | 0.8297 |
| KNN | 0.8933 | 0.9177 | 0.8934 | 0.8933 | 0.9422 | 0.9499 | 0.9577 | 0.9499 | 0.9499 | 0.9654 |
| LR | 0.5140 | 0.5480 | 0.4259 | 0.5140 | 0.5820 | 0.6682 | 0.6514 | 0.6278 | 0.6682 | 0.6346 |
| MLP | 0.7928 | 0.8265 | 0.7879 | 0.7928 | 0.8603 | 0.9090 | 0.9137 | 0.9079 | 0.9090 | 0.9184 |
| NB | 0.4854 | 0.5508 | 0.4412 | 0.4854 | 0.6162 | 0.2120 | 0.5038 | 0.1777 | 0.2120 | 0.7955 |
| RF | 0.9106 | 0.9327 | 0.9109 | 0.9106 | 0.9547 | 0.9594 | 0.9664 | 0.9594 | 0.9594 | 0.9734 |
| SVM | 0.3435 | 0.5195 | 0.3076 | 0.3435 | 0.6956 | 0.4634 | 0.6168 | 0.4819 | 0.4634 | 0.7702 |
| XGB | 0.7224 | 0.7655 | 0.7104 | 0.7224 | 0.8087 | 0.8467 | 0.8458 | 0.8381 | 0.8467 | 0.8449 |

**Table 5:** Experimental Result of Dataset-4

| Classifier | Traditional Approach |  |  |  |  | TClustVID |  |  |  |  |
| --- | --- | --- | --- | --- | --- | --- | --- | --- | --- | --- |
|  | Accuracy | AUC | F-Measure | Sensitivity | Specificity | Accuracy | AUC | F-Measure | Sensitivity | Specificity |
| DT | 0.8924 | 0.9148 | 0.8923 | 0.8924 | 0.9371 | 0.9428 | 0.9494 | 0.9426 | 0.9428 | 0.9560 |
| GB | 0.6211 | 0.6138 | 0.5530 | 0.6211 | 0.6065 | 0.8183 | 0.7880 | 0.8060 | 0.8183 | 0.7578 |
| KNN | 0.8727 | 0.9010 | 0.8729 | 0.8727 | 0.9292 | 0.9302 | 0.9392 | 0.9302 | 0.9302 | 0.9482 |
| LR | 0.5466 | 0.5556 | 0.4565 | 0.5466 | 0.5646 | 0.7472 | 0.7223 | 0.7279 | 0.7472 | 0.6974 |
| MLP | 0.7648 | 0.7969 | 0.7578 | 0.7648 | 0.8290 | 0.8817 | 0.8768 | 0.8781 | 0.8817 | 0.8719 |
| NB | 0.5331 | 0.5360 | 0.4218 | 0.5331 | 0.5389 | 0.2739 | 0.5055 | 0.1390 | 0.2739 | 0.7371 |
| RF | 0.8919 | 0.9176 | 0.8923 | 0.8919 | 0.9432 | 0.9425 | 0.9500 | 0.9424 | 0.9425 | 0.9575 |
| SVM | 0.3966 | 0.5187 | 0.3983 | 0.3966 | 0.6409 | 0.3257 | 0.5229 | 0.3398 | 0.3257 | 0.7201 |
| XGB | 0.6827 | 0.6907 | 0.6478 | 0.6827 | 0.6987 | 0.8254 | 0.8087 | 0.8171 | 0.8254 | 0.7921 |

**Table 6:** Experimental Result of Dataset-5

| Classifier | Traditional Approach |  |  |  |  | TClustVID |  |  |  |  |
| --- | --- | --- | --- | --- | --- | --- | --- | --- | --- | --- |
|  | Accuracy | AUC | F-Measure | Sensitivity | Specificity | Accuracy | AUC | F-Measure | Sensitivity | Specificity |
| DT | 0.8660 | 0.8991 | 0.8659 | 0.8660 | 0.9323 | 0.9017 | 0.9254 | 0.9017 | 0.9017 | 0.9490 |
| GB | 0.5341 | 0.6245 | 0.4940 | 0.5341 | 0.7149 | 0.6237 | 0.6839 | 0.5873 | 0.6237 | 0.7441 |
| KNN | 0.8406 | 0.8803 | 0.8406 | 0.8406 | 0.9201 | 0.8781 | 0.9073 | 0.8780 | 0.8781 | 0.9366 |
| LR | 0.4309 | 0.5519 | 0.3667 | 0.4309 | 0.6729 | 0.4543 | 0.5565 | 0.3862 | 0.4543 | 0.6586 |
| MLP | 0.6237 | 0.7123 | 0.6216 | 0.6237 | 0.8010 | 0.7485 | 0.7997 | 0.7437 | 0.7485 | 0.8510 |
| NB | 0.4189 | 0.5287 | 0.3053 | 0.4189 | 0.6385 | 0.4294 | 0.5244 | 0.3440 | 0.4294 | 0.6193 |
| RF | 0.8650 | 0.8997 | 0.8654 | 0.8650 | 0.9343 | 0.8995 | 0.9243 | 0.8996 | 0.8995 | 0.9492 |
| SVM | 0.3384 | 0.5248 | 0.2576 | 0.3384 | 0.7113 | 0.4239 | 0.5370 | 0.3617 | 0.4239 | 0.6502 |
| XGB | 0.5483 | 0.6469 | 0.5319 | 0.5483 | 0.7454 | 0.6447 | 0.7167 | 0.6390 | 0.6447 | 0.7886 |

**Table 7:** Experimental Result of Dataset-6

| Classifier | Traditional Approach |  |  |  |  | TClustVID |  |  |  |  |
| --- | --- | --- | --- | --- | --- | --- | --- | --- | --- | --- |
|  | Accuracy | AUC | F-Measure | Sensitivity | Specificity | Accuracy | AUC | F-Measure | Sensitivity | Specificity |
| DT | 0.8791 | 0.9085 | 0.8790 | 0.8791 | 0.9378 | 0.9324 | 0.9477 | 0.9324 | 0.9324 | 0.9629 |
| GB | 0.6021 | 0.6586 | 0.5615 | 0.6021 | 0.7151 | 0.7629 | 0.7847 | 0.7481 | 0.7629 | 0.8065 |
| KNN | 0.8584 | 0.8934 | 0.8585 | 0.8584 | 0.9285 | 0.9170 | 0.9359 | 0.9170 | 0.9170 | 0.9548 |
| LR | 0.4744 | 0.5614 | 0.4003 | 0.4744 | 0.6484 | 0.5263 | 0.5811 | 0.4652 | 0.5263 | 0.6360 |
| MLP | 0.7139 | 0.7778 | 0.7117 | 0.7139 | 0.8417 | 0.8463 | 0.8740 | 0.8451 | 0.8463 | 0.9017 |
| NB | 0.4503 | 0.5222 | 0.3148 | 0.4503 | 0.5942 | 0.4750 | 0.5145 | 0.3280 | 0.4750 | 0.5540 |
| RF | 0.8788 | 0.9102 | 0.8793 | 0.8788 | 0.9415 | 0.9311 | 0.9475 | 0.9312 | 0.9311 | 0.9639 |
| SVM | 0.4176 | 0.5302 | 0.3408 | 0.4176 | 0.6428 | 0.5359 | 0.5678 | 0.4334 | 0.5359 | 0.5997 |
| XGB | 0.6415 | 0.7187 | 0.6372 | 0.6415 | 0.7959 | 0.7744 | 0.8129 | 0.7720 | 0.7744 | 0.8513 |

**Table 8:** Experimental Result of Dataset-7

| Classifier | Traditional Approach |  |  |  |  | TClustVID |  |  |  |  |
| --- | --- | --- | --- | --- | --- | --- | --- | --- | --- | --- |
|  | Accuracy | AUC | F-Measure | Sensitivity | Specificity | Accuracy | AUC | F-Measure | Sensitivity | Specificity |
| DT | 0.9078 | 0.9294 | 0.9078 | 0.9078 | 0.9511 | 0.9550 | 0.9647 | 0.9550 | 0.9550 | 0.9745 |
| GB | 0.6637 | 0.7184 | 0.6555 | 0.6637 | 0.7731 | 0.8096 | 0.8298 | 0.8062 | 0.8096 | 0.8499 |
| KNN | 0.8886 | 0.9152 | 0.8887 | 0.8886 | 0.9417 | 0.9412 | 0.9540 | 0.9412 | 0.9412 | 0.9667 |
| LR | 0.4511 | 0.5376 | 0.3804 | 0.4511 | 0.6242 | 0.5484 | 0.5978 | 0.5007 | 0.5484 | 0.6473 |
| MLP | 0.7681 | 0.8133 | 0.7644 | 0.7681 | 0.8585 | 0.8854 | 0.9054 | 0.8850 | 0.8854 | 0.9253 |
| NB | 0.2189 | 0.5008 | 0.0825 | 0.2189 | 0.7826 | 0.2199 | 0.5033 | 0.0944 | 0.2199 | 0.7868 |
| RF | 0.9085 | 0.9313 | 0.9087 | 0.9085 | 0.9542 | 0.9540 | 0.9644 | 0.9541 | 0.9540 | 0.9748 |
| SVM | 0.3525 | 0.5166 | 0.3527 | 0.3525 | 0.6808 | 0.2991 | 0.5394 | 0.2507 | 0.2991 | 0.7797 |
| XGB | 0.6354 | 0.7047 | 0.6322 | 0.6354 | 0.7741 | 0.8150 | 0.8431 | 0.8142 | 0.8150 | 0.8711 |

**Table 9:**
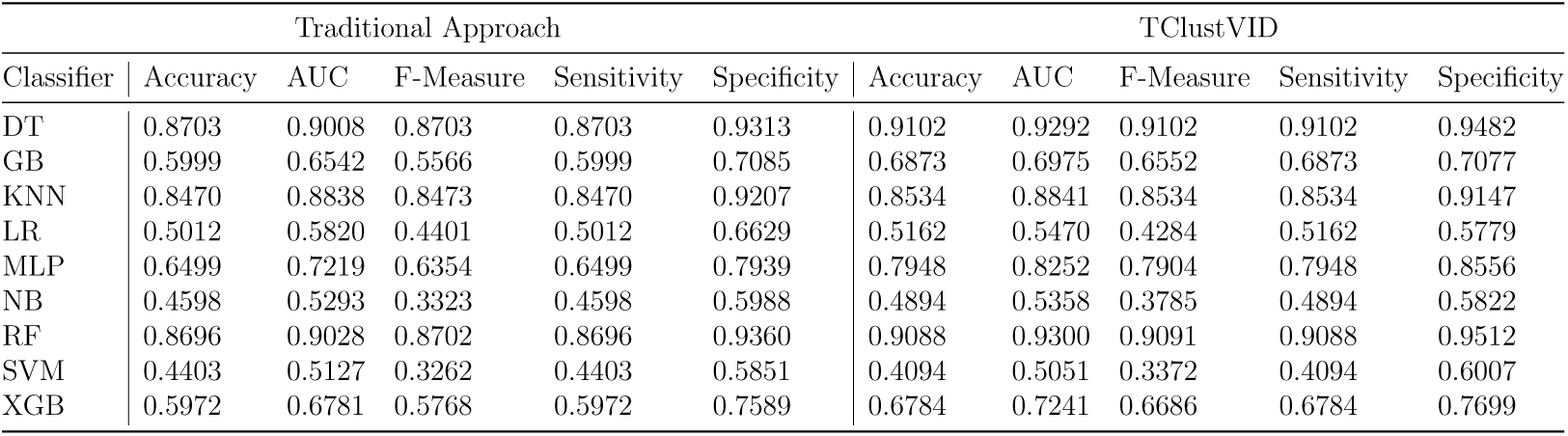
Experimental Result of Dataset-8

**Table 10:** Experimental Result of Dataset-9

| Classifier | Traditional Approach |  |  |  |  | TClustVID |  |  |  |  |
| --- | --- | --- | --- | --- | --- | --- | --- | --- | --- | --- |
|  | Accuracy | AUC | F-Measure | Sensitivity | Specificity | Accuracy | AUC | F-Measure | Sensitivity | Specificity |
| DT | 0.8697 | 0.9004 | 0.8697 | 0.8697 | 0.9310 | 0.9111 | 0.9301 | 0.9111 | 0.9111 | 0.9490 |
| GB | 0.6000 | 0.6544 | 0.5568 | 0.6000 | 0.7089 | 0.6860 | 0.6983 | 0.6513 | 0.6860 | 0.7105 |
| KNN | 0.8467 | 0.8837 | 0.8470 | 0.8467 | 0.9206 | 0.8557 | 0.8863 | 0.8558 | 0.8557 | 0.9168 |
| LR | 0.4981 | 0.5790 | 0.4365 | 0.4981 | 0.6599 | 0.5077 | 0.5409 | 0.4200 | 0.5077 | 0.5741 |
| MLP | 0.6503 | 0.7153 | 0.6331 | 0.6503 | 0.7803 | 0.8018 | 0.8304 | 0.7970 | 0.8018 | 0.8590 |
| NB | 0.2206 | 0.5003 | 0.0829 | 0.2206 | 0.7800 | 0.2504 | 0.5072 | 0.1914 | 0.2504 | 0.7641 |
| RF | 0.8691 | 0.9024 | 0.8697 | 0.8691 | 0.9357 | 0.9101 | 0.9312 | 0.9104 | 0.9101 | 0.9523 |
| SVM | 0.3449 | 0.5081 | 0.2999 | 0.3449 | 0.6712 | 0.2698 | 0.5145 | 0.2426 | 0.2698 | 0.7592 |
| XGB | 0.5992 | 0.6796 | 0.5785 | 0.5992 | 0.7600 | 0.6762 | 0.7259 | 0.6675 | 0.6762 | 0.7755 |

In this work, we implemented TClustVID where these results of individual classifiers have been improved over the traditional approaches (see Table-2, 3, 4, 5, 6,7, 8, 9 and 10). However, the performance order of individual classification remained almost the same. Various classifiers such as DT, RF, GB, KNN, MLP, RF, SVM and XGB were employed to compare with TClustVID. Moreover, the performance of DT and RF were the most similar when investigating COVID-19 twitter data analysis for various evaluation metrics. Some dissimilarities were noted when the data processing methodology is modified in different steps. Using the traditional approaches, RF showed better results than DT except accuracy and sensitivity. When TClustVID was employed, the performance of DT increased comparing to the traditional approach. DT showed 7,6,7,7 and 2 times the highest accuracy, AUC, f-measure, sensitivity and specificity at the nine COVID-19 twitter datasets respectively. Again, RF showed 2,3,2,2 and 7 times the highest accuracy, AUC, f-measure, sensitivity and specificity. The results of KNN and MLP were improved but were still third and fourth high performing classifiers for all of these datasets. With the traditional approach, GB showed better performance than XGB under various conditions. However, the results of XGB showed superiority to GB in almost all the time with TClustVID, so that GB showed greater accuracy and sensitivity than XGB in dataset-8 and 9. Moreover, LR and SVM showed lower performance than GB whereas LR showed better performance than SVM.

However, the average results for the combination of traditional and TClustVID are illustrated in Figure 1). Subsequently, the individual average results of traditional and TClustVID were explored to understand the average hierarchy of individual classifiers for both of these approaches. Using Traditional approach, RF showed 7 times and DT showed 2 times top results corresponding all metrics respectively. Thus, RF considered the best performing and DT represented the second best performing classifier in this analysis, with KNN and MLP third and fourth classifier in terms of their performance. Besides, XGB, GB, LR and SVM showed 8 times as the fifth, sixth, seventh best performing algorithms respectively. Instead, the average results of almost all classifiers were improved by TClustVID. DT showed greater average result with TClustVID, where it showed the highest outcomes at the five twitter clusters. Thus, RF can be considered as the second highest average performing classifier in this work. KNN and MLP showed the third and fourth highest performing classifier in both of these approaches. Therefore, XGB, GB, LR and SVM that showed the next best average performance.

**Figure 1:**
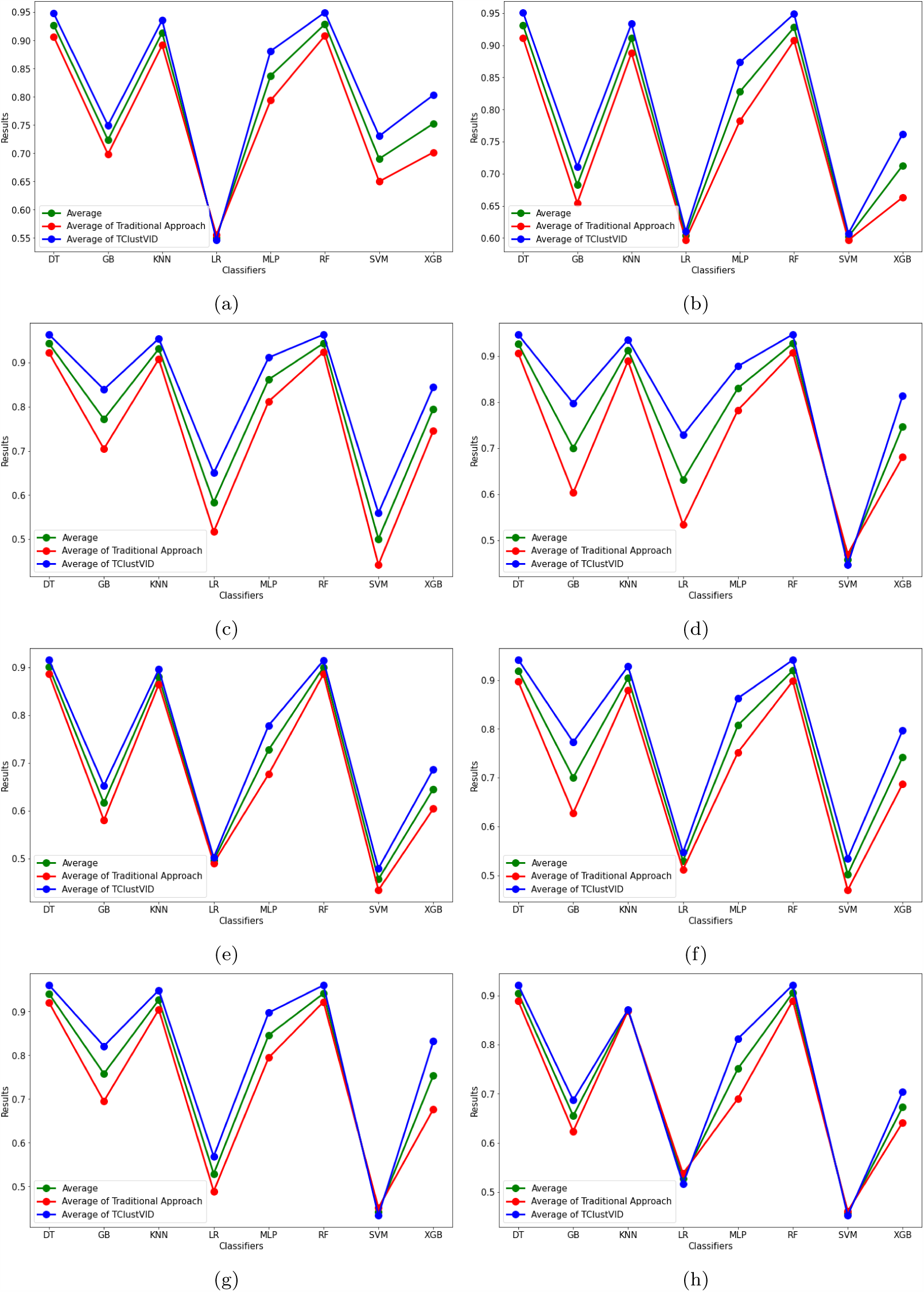
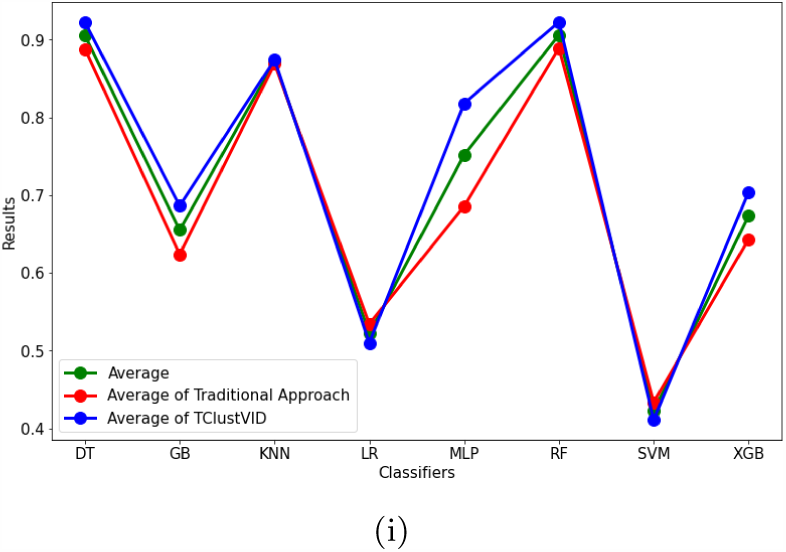
Average Performance of various classifiers in (a) Dataset-1, (b) Dataset-2, (c) Dataset-3, (d) Dataset-4, (e) Dataset-5, (f) Dataset-6, (g) Dataset-7, (h) Dataset-8 and (i) Dataset-9 of traditional and TClustVID

The highest results of different classifiers are indicated the best performance in analyzing COVID-19 tweets. Therefore, the highest results for different classifiers are shown on Table 11. In this table, the findings of TClustVID were also shown improved outcomes relative to the traditional approach. In both apporaches, RF showed the best results among all of the classification methods. Then, DT showed the second maximum results to investigate COVID-19 related tweets. Again, KNN and MLP showed the third and fourth best results, similar to previous analyses. We then found that XGB and GB also gave better results, with XGB giving better results than GB. Using the traditional approach, SVM showed greater accuracy, AUC, sensitivity and specificity than LR. Instead, LR showed greater accuracy, AUC and sensitivity in TClustVID.

**Table 11:** Highest Results of Different Classifiers

| Classifier | Traditional Approach |  |  |  |  | TClustVID |  |  |  |  |
| --- | --- | --- | --- | --- | --- | --- | --- | --- | --- | --- |
|  | Accuracy | Auc | F-Measure | Sensitivity | Specificity | Accuracy | Auc | F-Measure | Sensitivity | Specificity |
| DT | 0.9309 | 0.9304 | 0.9310 | 0.9309 | 0.9511 | 0.9637 | 0.9666 | 0.9637 | 0.9637 | 0.9745 |
| GB | 0.8158 | 0.7184 | 0.7550 | 0.8158 | 0.7731 | 0.8563 | 0.8380 | 0.8360 | 0.8563 | 0.8499 |
| KNN | 0.9238 | 0.9177 | 0.9220 | 0.9238 | 0.9422 | 0.9575 | 0.9577 | 0.9569 | 0.9575 | 0.9667 |
| LR | 0.7866 | 0.5820 | 0.6953 | 0.7866 | 0.6729 | 0.8072 | 0.7223 | 0.7279 | 0.8072 | 0.6974 |
| MLP | 0.8673 | 0.8265 | 0.8543 | 0.8673 | 0.8603 | 0.9245 | 0.9137 | 0.9212 | 0.9245 | 0.9253 |
| RF | 0.9374 | 0.9327 | 0.9361 | 0.9374 | 0.9547 | 0.9676 | 0.9664 | 0.9672 | 0.9676 | 0.9748 |
| SVM | 0.7867 | 0.5996 | 0.6947 | 0.7867 | 0.7113 | 0.8063 | 0.6928 | 0.7718 | 0.8063 | 0.7797 |
| XGB | 0.8196 | 0.7655 | 0.7635 | 0.8196 | 0.8087 | 0.8745 | 0.8458 | 0.8547 | 0.8745 | 0.8711 |
| AVG | 0.8585 | 0.7841 | 0.8190 | 0.8585 | 0.8343 | 0.8947 | 0.8629 | 0.8749 | 0.8947 | 0.8799 |

After calculating the average results of the different classifiers, it was clear that TClustVID showed better results compared to the more traditional approach (see Table 12). However, the order of average performances is similar to whether the traditional approach or TClustVID was used. RF showed the highest average accuracy, f-measure and sensitivity and was the highest average classification model in this analysis. Instead, DT appears as the second ranked for average performing classifier and KNN and MLP were found third and fourth performing classifier, which was also seen in another analysis. XGB showed a better average performance than GB. Hence, XGB and GB represent the fifth and sixth performing classifiers in this work. Finally, LR and SVM show the lowest average order of performance.

**Table 12:** Average Results of Different Classifiers

| Classifier | Traditional Approach |  |  |  |  | TClustVID |  |  |  |  |
| --- | --- | --- | --- | --- | --- | --- | --- | --- | --- | --- |
|  | Accuracy | Auc | F-Measure | Sensitivity | Specificity | Accuracy | Auc | F-Measure | Sensitivity | Specificity |
| DT | 0.8936 | 0.9092 | 0.8936 | 0.8936 | 0.9248 | 0.9365 | 0.9445 | 0.9365 | 0.9365 | 0.9524 |
| GB | 0.6581 | 0.6511 | 0.6170 | 0.6581 | 0.6442 | 0.7674 | 0.7399 | 0.7444 | 0.7674 | 0.7125 |
| KNN | 0.8757 | 0.8911 | 0.8754 | 0.8757 | 0.9066 | 0.9143 | 0.9236 | 0.9142 | 0.9143 | 0.9328 |
| LR | 0.5443 | 0.5465 | 0.4642 | 0.5443 | 0.5488 | 0.6060 | 0.5781 | 0.5372 | 0.6060 | 0.5501 |
| MLP | 0.7412 | 0.7622 | 0.7329 | 0.7412 | 0.7832 | 0.8548 | 0.8594 | 0.8520 | 0.8548 | 0.8639 |
| RF | 0.8949 | 0.9092 | 0.8950 | 0.8949 | 0.9234 | 0.9367 | 0.9438 | 0.9367 | 0.9367 | 0.9510 |
| SVM | 0.4641 | 0.5257 | 0.4080 | 0.4641 | 0.5872 | 0.4818 | 0.5553 | 0.4381 | 0.4818 | 0.6289 |
| XGB | 0.6704 | 0.6799 | 0.6475 | 0.6704 | 0.6894 | 0.7765 | 0.7717 | 0.7679 | 0.7765 | 0.7668 |
| AVG | 0.7178 | 0.7344 | 0.6917 | 0.7178 | 0.7510 | 0.7843 | 0.7895 | 0.7659 | 0.7843 | 0.7948 |

In Fig-2a using the traditional approach, the sequence of average highest outcomes of different classifiers are also represented as RF, DT, KNN, MLP, XGB, GB, SVM and LR. Similarly, TClustVID represents the ranking of average best results of classifiers as RF, DT, KNN, MLP, XGB, GB, SVM and LR respectively. On the other hand, the average performance of averaged classification results is illustrated at Fig-2b. In the traditional approach, the sequences of average results of averaged classifiers are represented as RF, DT, KNN, MLP, XGB, GB, LR and SVM. However, the average performance of TClustVID, DT showed better results than RF. Moreover, the performance of other classifiers keep the same sequence according to the previous analysis. Thus, the final average order of top average performing classifiers in TClustVID were DT, RF, KNN, MLP, XGB, GB, LR and SVM. However, in the mixture of the traditional and TClustVID, average results showed a similar average ranking of performance of various classifiers which are RF, DT, KNN, MLP, XGB, GB, LR and SVM respectively.

Along with observing the performance of various classifiers, we noticed that TClustVID shows better performance than traditional approach. Hence, top modeling approach is used high performing clusters to extract significant topics in next section.

### 3.2 Topic Modeling Approach

A comprehensive analysis of different classifiers in traditional and TClustVID analyses indicated that TClustVID is the best model to identify significant groups of tweets from large COVID-19 Twitter datasets. The data obtained from identification of groups/clusters were significant because they showed the highest classification accuracy were achieved compared to traditional analysis in primary data. In the TClustVID analysis, we generated significant clusters from each of these twitter datasets (for positive neutral, and negative categories) that showed greatly improved results for the different classifiers. These clusters have been denoted as Cluster-1, Cluster-2, Cluster-3, Cluster-4, Cluster-5, Cluster-6, Cluster-7, Cluster-8, and Cluster-9, respectively. A number of topics were then extracted from these clusters where within nine clusters seven clusters produces positive, neutral and negative topics and two of them extracts positive and neutral topics using LDA. Each topic contains 10 tokens along with related weights and they can be used to prioritize each token. 20 topics were identified from each of the categories (positive, neutral and negative) in these clusters. Therefore, all topics of individual clusters are represented as word cloud in the supplementary section. In this paper, extracted positive, neutral and negative topics of cluster-3 are visualized with word cloud in Figure 4, 5 and 6 individually. However, LDA cannot interpret the meaning, so we defined each topic by realising the meaning and weight values at different groups manually. The positive, neutral and negative topics are represented at Table 13, 14 and 15 respectively. These tasks are not simple because many preprocessed words do not have any semantic meaning. However, it can be hard to understand the association between the different words/tokens in these topics and these interpretations may slightly differ with other types of reviews.

**Table 13:** Positive Topics of All Significant Clusters

|  | Cluster-1 | Cluster-2 | Cluster-3 | Cluster-4 | Cluster-5 |
| --- | --- | --- | --- | --- | --- |
| Topics-1 | Culture | Prevention | Kids | Wish | Sunny |
| Topics-2 | Nationality | Situation | Wish | News | Watch |
| Topics-3 | Prevention | Situation | Testing | Situation | Affect |
| Topics-4 | Caring | Homework | Treatment | Help | Situation |
| Topics-5 | Blaming | News | Testing | Help | Treatment |
| Topics-6 | Believe | News | Caring | Facts | Awareness |
| Topics-7 | Die | News | Feeling | Control | Medicine |
| Topics-8 | Caring | Wish | Situation | Infectious | Treatment |
| Topics-9 | Discrimination | Awareness | Scaring | Right | Medicine |
| Topics-10 | Situation | Financial State | Buying | Awareness | Awareness |
| Topics-11 | Crisis | News | Fun | Wish | Prevention |
| Topics-12 | Financial Help | Avoidness | Right | News | Situation |
| Topics-13 | Condition | Crisis | Panic | Situation | Awareness |
| Topics-14 | Wish | Food | Protection | Distance & Treatment | Treatment |
| Topics-15 | Lockdown | Blaming | Health | Annoying | Awareness |
| Topics-16 | Closing | Situation | Awareness | Situation | Humor |
| Topics-17 | Closing | Lockdown | Panic | Job | Situation |
| Topics-18 | Awareness | Awareness | Effect | Stay Safe | Risk |
| Topics-19 | Financial Help | Annoying | Micro-Organism | Awareness | Situation |
| Topics-20 | Caring | Awareness | News | Wish | Risk |
|  | Cluster-6 | Cluster-7 | Cluster-8 | Cluster-9 |  |
| Topics-1 | Right | Testing & Treatment | Survive | Shut |  |
| Topics-2 | Need | Interest | Flu | Honest |  |
| Topics-3 | Covid | Need | Move | Media |  |
| Topics-4 | Social Media | Social Distance | Overreact | Right |  |
| Topics-5 | Awareness | Social Distance | Situation | Testing |  |
| Topics-6 | Flight | Epidemic | Rumor | Caring |  |
| Topics-7 | Messege | Social Distance | Fight & Caring | Isolation |  |
| Topics-8 | Right | Symptoms | Cases | Survive |  |
| Topics-9 | Treatment | Effect | Disease | Home |  |
| Topics-10 | Wish | Confirmed | Cases | Wish |  |
| Topics-11 | Situation | Coronavirus | Awareness | Worried |  |
| Topics-12 | Warning | Message | Infectious | Situation |  |
| Topics-13 | Testing & Treatment | Coronavirus | Social Guys | Quarantine |  |
| Topics-14 | Cases | Social Distance | Situation | Love |  |
| Topics-15 | Message | Tourism | Quarentine | Scaring |  |
| Topics-16 | Message | Tourism | Awareness | Don't Move |  |
| Topics-17 | Situation | Coronavirus | Facts | Affect |  |
| Topics-18 | Tourism | Outbreak | Schools | Wind |  |
| Topics-19 | Coronavirus | Coronavirus | Crisis & Prevention | Awareness |  |
| Topics-20 | Awareness | Awareness | Financial Enrichment | Fuck |  |

**Table 14:** Neutral Topics of All Significant Clusters

|  | Cluster-1 | Cluster-2 | Cluster-3 | Cluster-4 | Cluster-5 |
| --- | --- | --- | --- | --- | --- |
| Topics-1 | Financial Lose | Warning | Outbreak | Situation | Awareness |
| Topics-2 | Fact | Food | Sharing | Panic | Infectious |
| Topics-3 | Warning | Situation | Wish | Situation | Situation |
| Topics-4 | Estimate | Situation | Gonna | Entertainment | Need |
| Topics-5 | Blaming | Testing | Caring | Protection | Wish |
| Topics-6 | Pleased | Rumor | Caring | Dead | Food |
| Topics-7 | Financial Lose | Warning | Panic | Health | Break |
| Topics-8 | Pandemic Warning | Visiting | Survive | Stay Home | Treatment |
| Topics-9 | Awareness | Joke | Awareness | Avoid | Want |
| Topics-10 | Disease | Panic | Treatment | Fact | Prevention |
| Topics-11 | Warning | Situation | Playing Game | Awareness | Awareness |
| Topics-12 | Caring | Panic | Coronavirus | Protection | Panic |
| Topics-13 | Panic | Closing | Homework | Awareness | Situation |
| Topics-14 | Panic | Panic | Ramadhan News | Situation | Awareness |
| Topics-15 | Awareness | Panic | Sanitation | Fact | Prevention |
| Topics-16 | Panic | Situation | Wish | Panic | Coronavirus |
| Topics-17 | Blaming | Homework | Situation | Wish | Avoid |
| Topics-18 | Joke | Blaming | Coronavirus | Update | Food |
| Topics-19 | Joke | Panic | Avoid | Cases | Situation |
| Topics-20 | Annoyed | Annoyed | Stop Spreading | Hospitalize | Coronavirus |
|  | Cluster-6 | Cluster-7 | Cluster-8 | Cluster-9 |  |
| Topics-1 | Vacine | Ruin | Situation | Tourism |  |
| Topics-2 | News | Cases | Watch | Outbreak |  |
| Topics-3 | Message | Coronavirus | Virus | Situation |  |
| Topics-4 | Prevention | Awareness | Touch | Situation |  |
| Topics-5 | Dead | Wait & Things | Symptom | Quarantine |  |
| Topics-6 | News | Crisis | Problem | Education |  |
| Topics-7 | Panic | Symptom | Shot | Education |  |
| Topics-8 | Protection | News | Like | Virus |  |
| Topics-9 | Awareness | Symptom | Situation | Pandemic |  |
| Topics-10 | Situation | Infectious | Sick | Dead |  |
| Topics-11 | Thread | Expose | Dead | Education |  |
| Topics-12 | Wish | Caring | Body | Awareness |  |
| Topics-13 | Situation | Help & Need | Flu | Body |  |
| Topics-14 | Awareness | Protection | Wish | Need |  |
| Topics-15 | Message | Testing | Panic | Caring |  |
| Topics-16 | Situation | Blaming | Watch | Panic |  |
| Topics-17 | Media | Cure | Time | Fact |  |
| Topics-18 | Coronavirus | Message | Panic | Cases |  |
| Topics-19 | Cases | Stay Home | Contract | Public |  |
| Topics-20 | Health | Situation | Awareness | Exhibit |  |

**Table 15:**
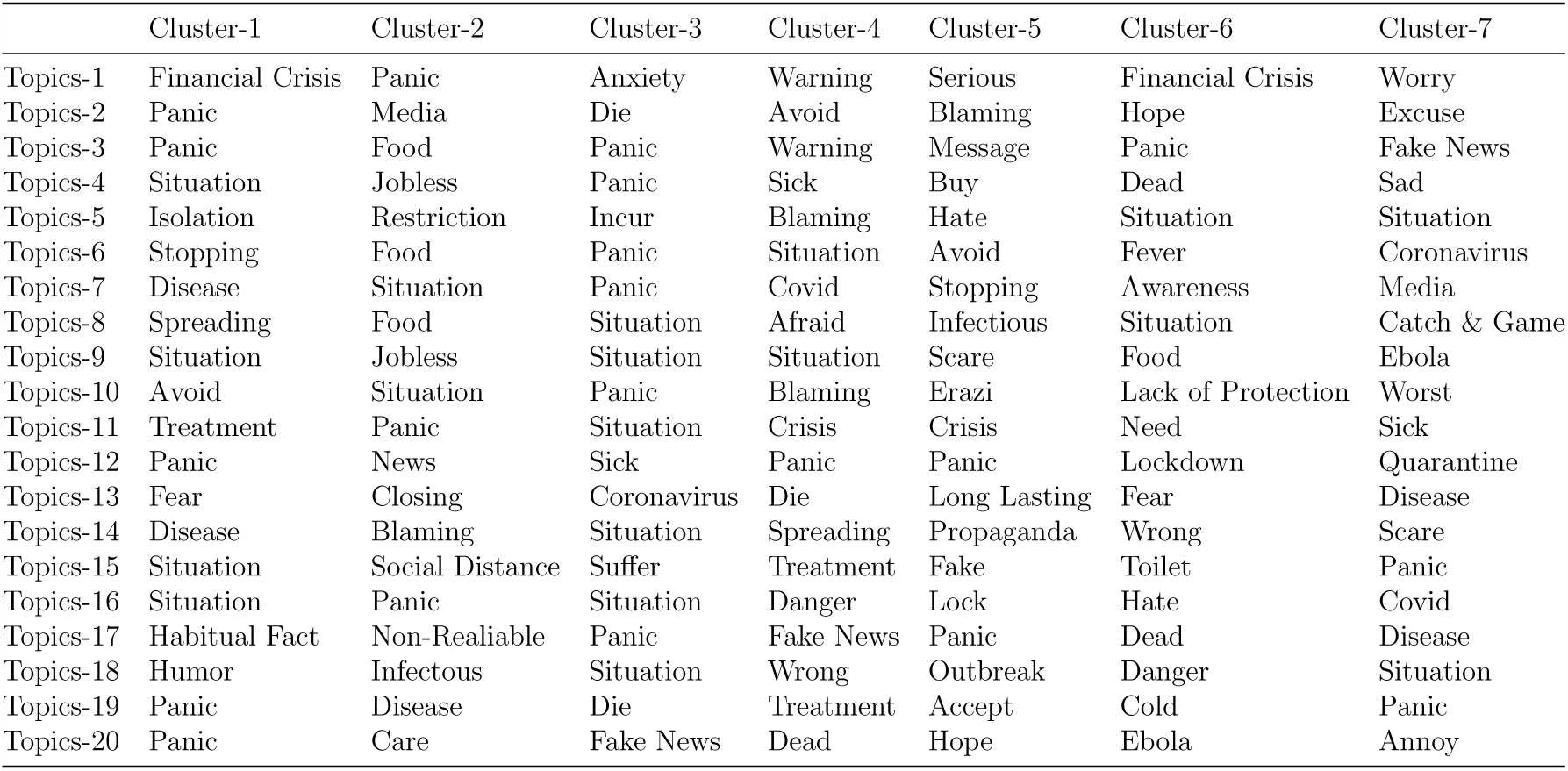
Negative Topics of All Significant Clusters

**Figure 2:**
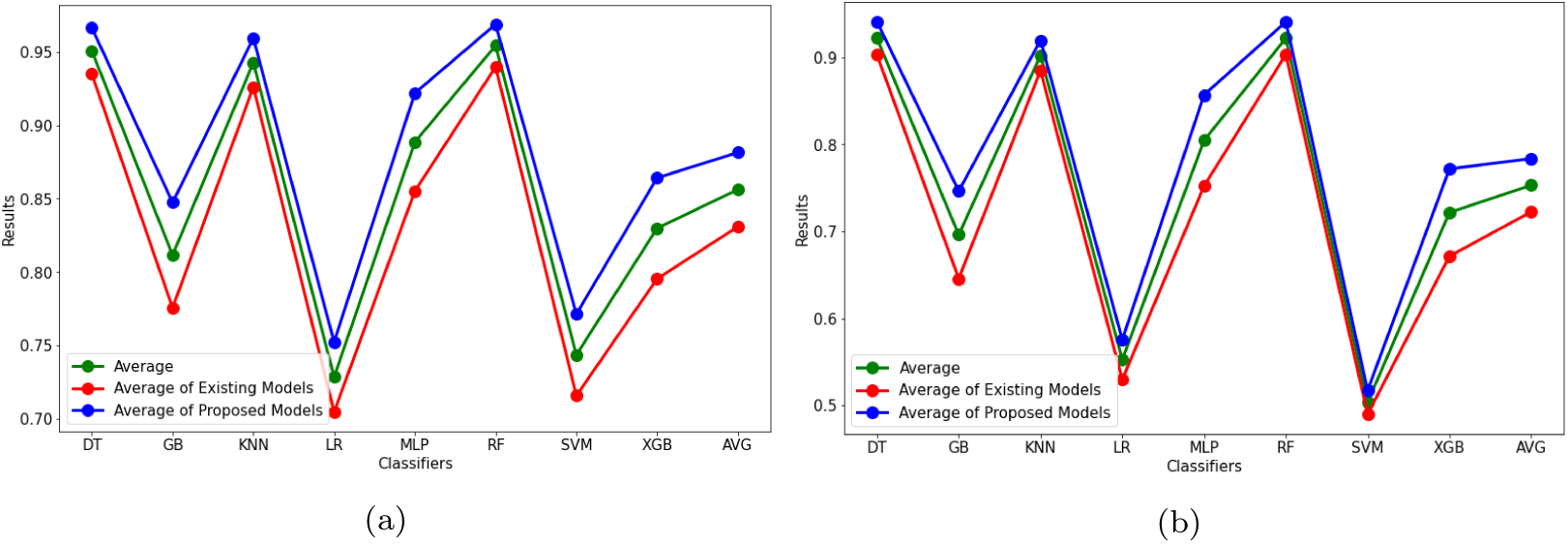
Average Performance of various classifiers (a) Maximum results (b) Average results corresponding to the nine twitter experimental datasets

**Figure 3:**
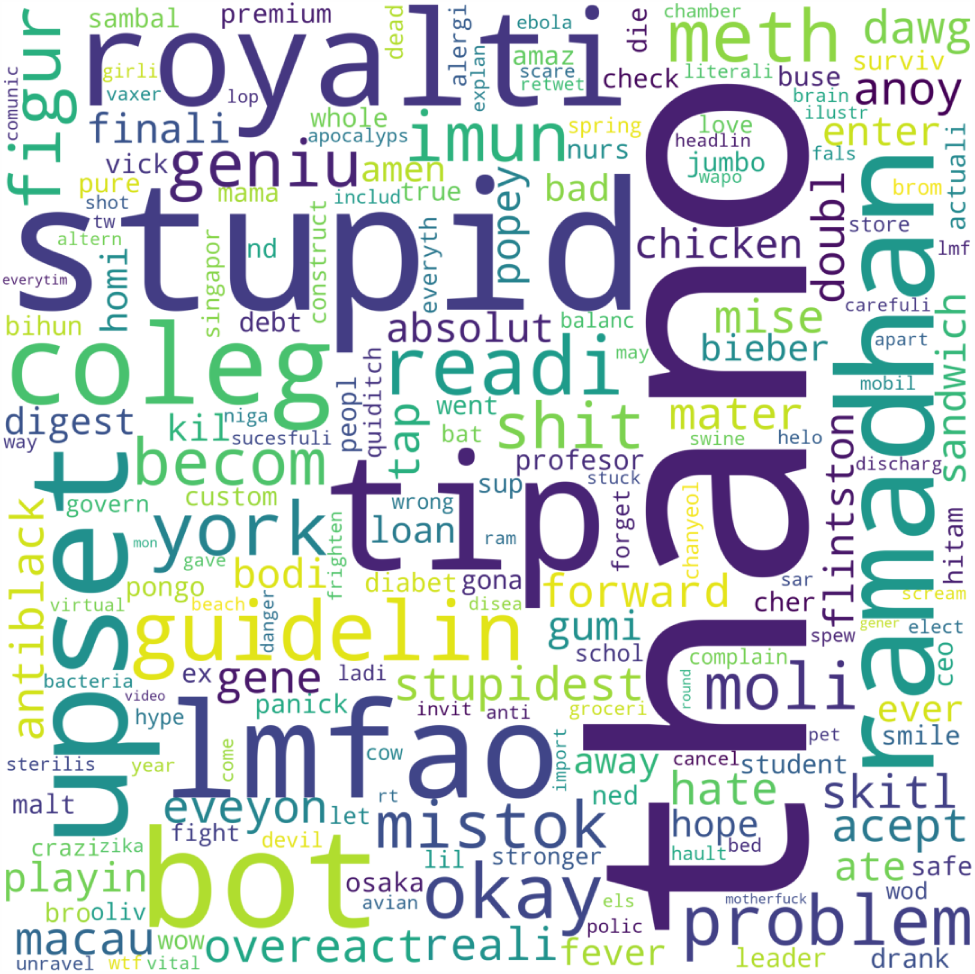
Word Cloud of Various Topics

**Figure 4:**
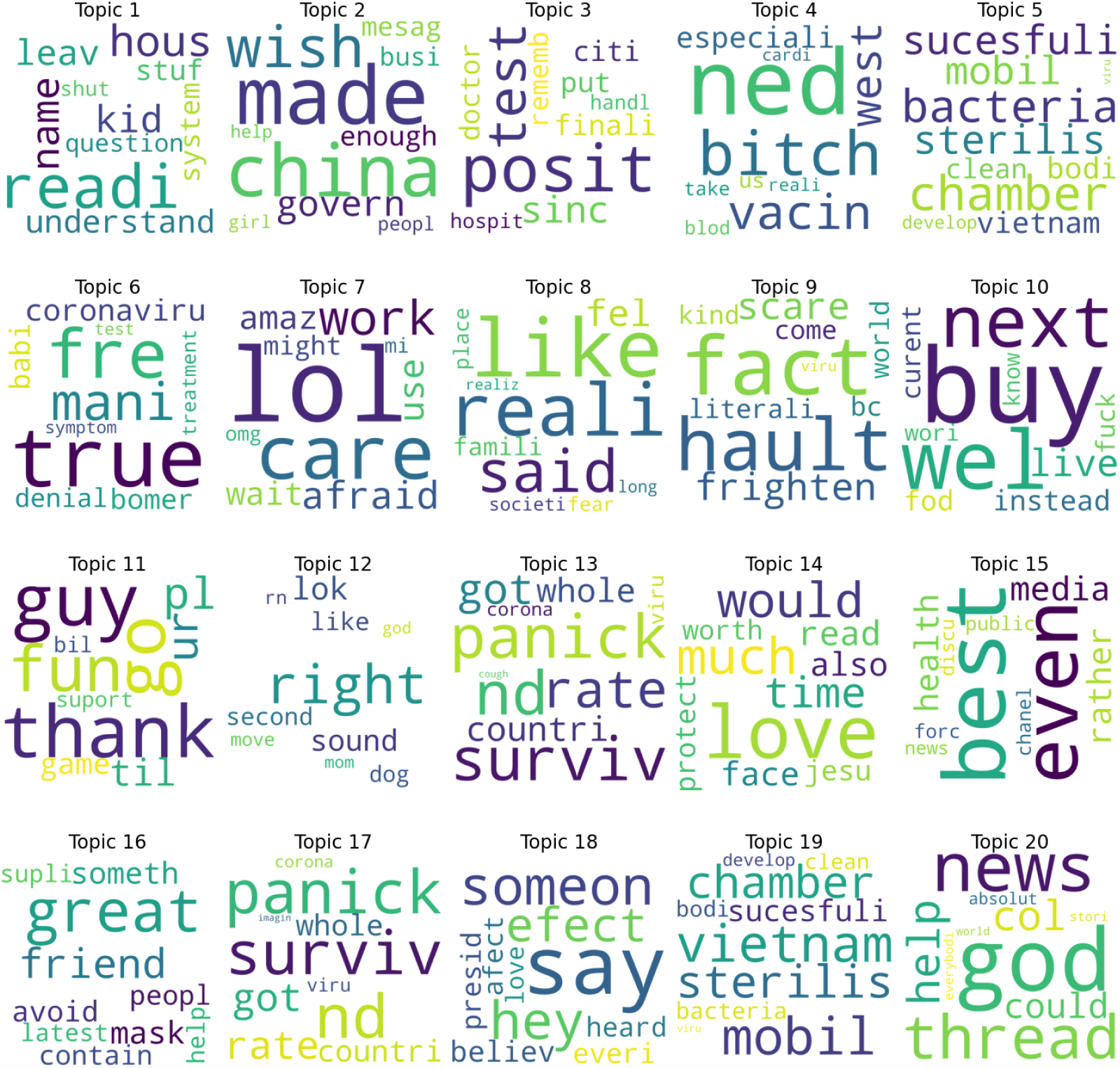
Positive Topics of Cluster-3

**Figure 5:**
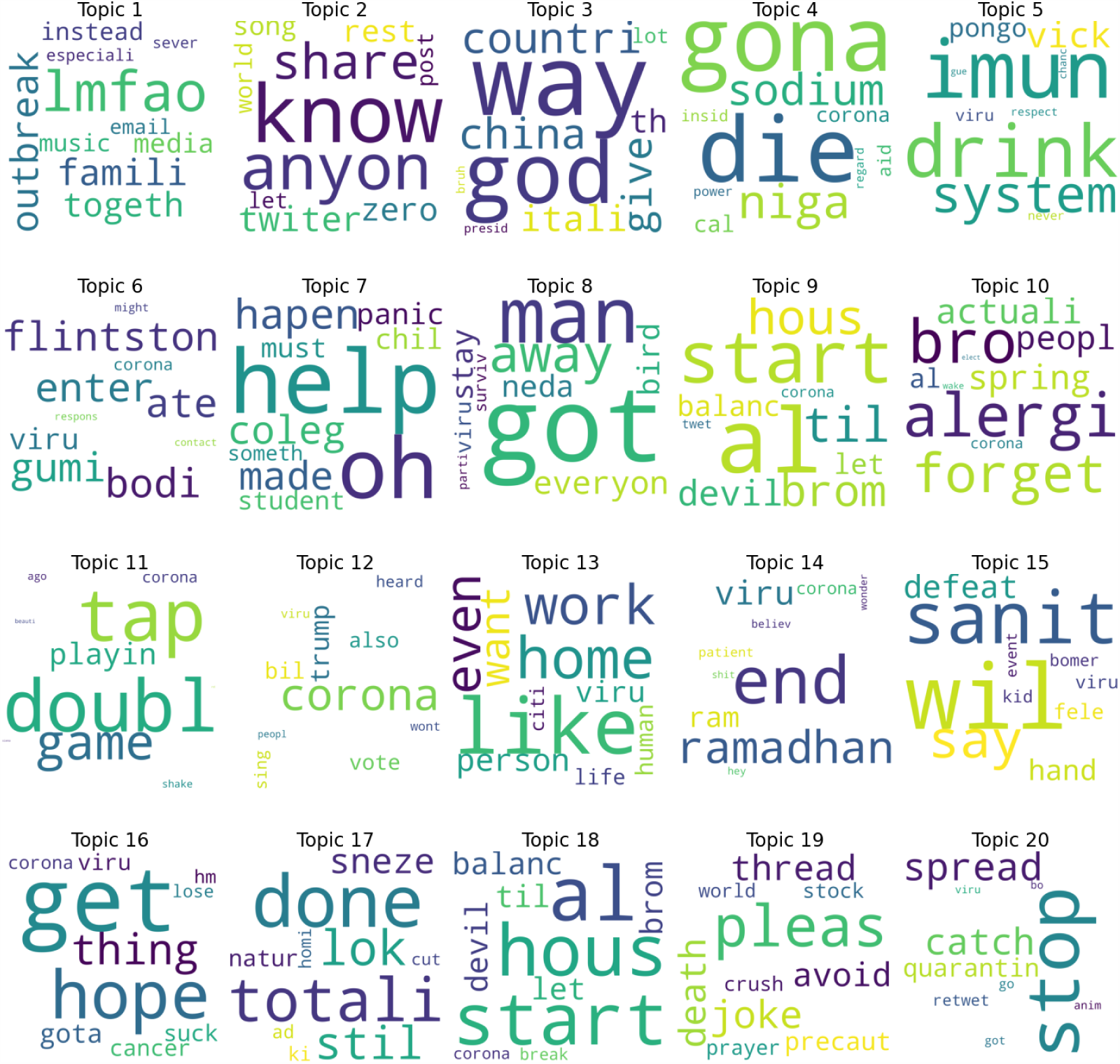
Neutral Topics of Cluster-3

**Figure 6:**
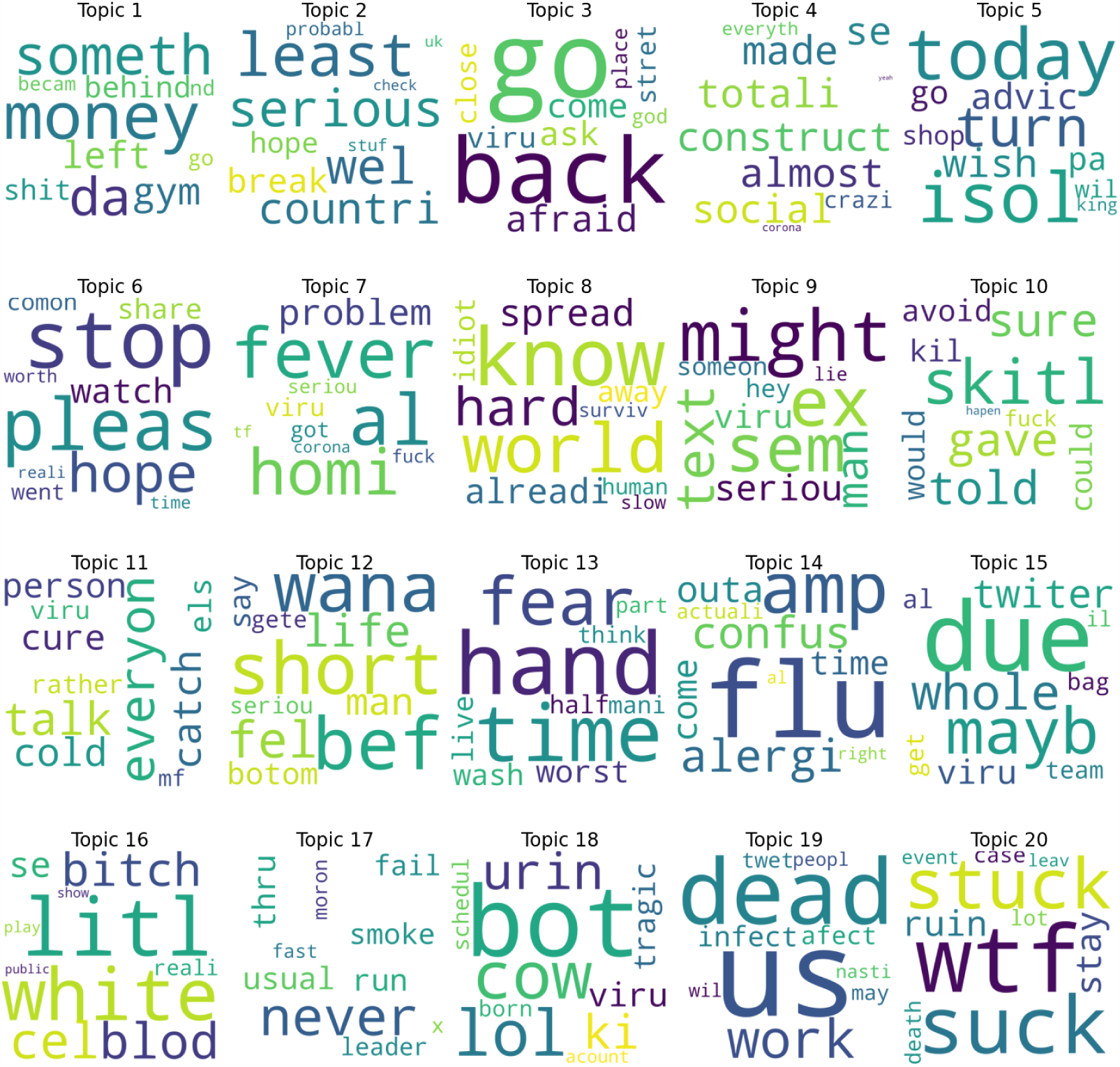
Negative Topics of Cluster-3

In the different categories of tweets, we manipulated the frequency of different topics that appears several times. Positive, neutral and negative topics were represented to identify what activities are generated in the context. To understand individual topics into different categories, we considered the best topics which are appeared more than 1 times (see Figure 7). The examples of positive topics of cluster-3 are shown as the word cloud in Figure 4. The positive topics of different clusters are shown in Table 13 and the top frequent positive topics are shown in Figure 7a. For the positive cases, ‘awareness’ and ‘situation’ are the most frequent topics that appear many times in different clusters. Both of these appear 17 times in different significant clusters. ‘Awareness’ is specified those actions whose are taken by individuals and situation symbolizes the general situation of particular places/incidents where pandemic news indicates a generic situation relating to COVID-19. ‘Wishes’ appear 8 and ‘new’ appears 7 times in this study. Furthermore, ‘caring’, ‘coronavirus’, ‘right’ and ‘treatment’ are found 5 times, and ‘message’, and ‘social distance’ are found 4 times this effort. Subsequently, ‘cases’, ‘prevention’, ‘testing’ and ‘tourism’ are found 3 times in the COVID-19 situation. In addition, other precaution related topics such as ‘affect’, ‘annoying’, ‘blaming’, ‘closing’, ‘crisis’, ‘effect’, ‘facts’, ‘financial help’, ‘help’, ‘infectious’, ‘lockdown’, ‘medicine’, ‘need’, ‘panic’, ‘quarantine’, ‘risk’ and ‘scaring’ are shown their frequency 2 times in different clusters. These are appeared regularly and specifies how we can improve this condition. However, some of negative topics, for instance’blaming’,’crisis’,’infectious’, ‘panic’, ‘risk’ appeared in positive cases but their frequencies are not greater. More upcoming positive issues are also addressed in this analysis included ‘financial help, ‘, ‘help’, ‘lockdown’, ‘quarantine’ and ‘medicine.

**Figure 7:**
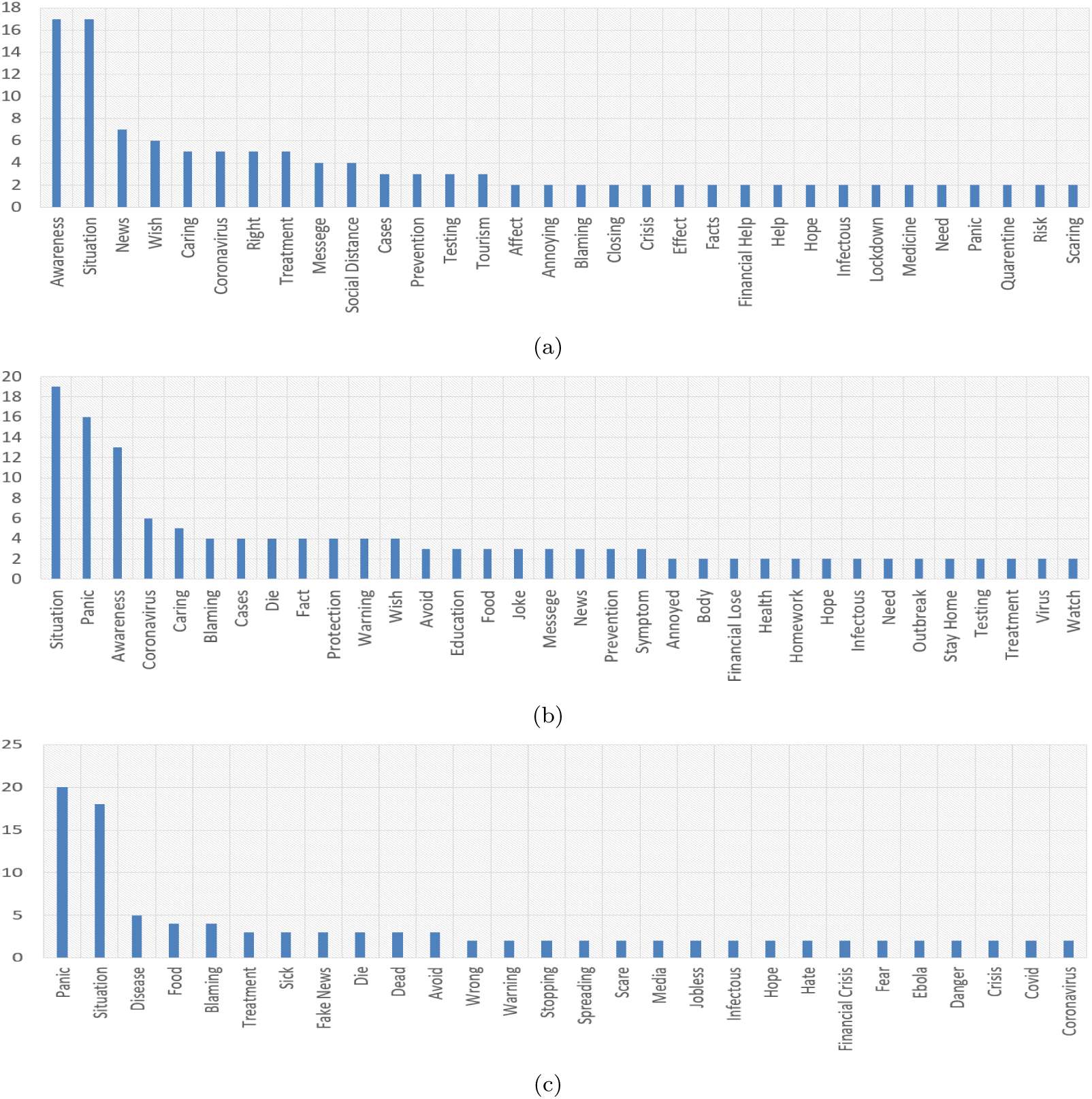
Top Frequency of (a) Positive (b) Neutral (c) Negative COVID-19 Associated Topics

In the neutral category, there are appeared the mixture of positive and negative topics which indicates the most frequent topics in recent times. For example, we represent an example of neutral topics as a world cloud is shown in Figure 5. Besides this, neutral topics of different clusters are provided in Table 14 and top frequent topics are shown at Figure 7b. Therefore, ‘situation’, ‘panic’ and ‘awareness’ are found 19, 16 and 13 times in the following list of twitter topics. ‘Panic’ is a related topic to explain epidemic conditions and news. In addition, ‘wish’ and ‘coronavirus’ appear 6 times as well as ‘caring’ which appears 5 times at negative tweets. Consequently, ‘blaming’, ‘cases’, ‘die’, ‘warning’ and ‘protection’ appear 4 times while education, ‘food’, ‘joke’, ‘message’, ‘news’, ‘prevention’, and ‘symptom’ appear 3 times in this condition. The rest of the topics perform 2 times to represent as neutral topics.

The more upcoming issue before and after COVID-19like’Financial’,’lose’,’crisis’,’food’, ‘education’ also arose in this analysis.

The negative topics using the word cloud are represented in Figure 6. Thus, the meaning of negative topics has been provided in Table 15 and topmost frequent topics are shown in Figure 7c. In this category, panic and situation appear most of the times than other topics. Both of them appear 20 and 18 times respectively. ‘Dead’ and ‘disease’ appear 6 and 5 times enabling estimation of its influence. Thus, ‘food’ and ‘blaming’ appear 4 times and ‘treatment’, ‘sick’, ‘fake news’ and ‘avoid’ appear 3 times to represent significant topics. Some cases like ‘food’ and ‘treatment’ indicate the level of crisis perceived. The rest of the topics presented with a frequency of 2 in this work. Therefore, these topics shown in the top list indicate feelings or perceptions relating to the COVID-19 that are negative.

### 3.3 Implication

Therefore, we explored different topics that represent feelings or perceptions that relate to the current situation of the COVID-19 pandemic. Every day many people share their idea, opinion, argument etc. on different social media like twitter. But, these huge amount of opinion cannot represent to the general people more frequently. In this case, this dynamic topics modeling is so much helpful to understand this pandemic and predict the future condition. Proposed TClustVID shows more accuracy than traditional approach. In high performing clusters, we extracted positive, neutral and negative topics to investigate what mattered to the tweets and realized the associated topics of individual categories. These opinions and comments on social media reflecting significant values and gives various information about related issues. Hence, these topics can be informative to government and policymakers that need to make a rapid decision and deal with the uncertain COVID-19 situation using the best available information. In addition, these types of analysis help to clarify the concerns to the people finding themselves experiencing the pandemic situation in every day. The most frequently raised topics thus indicate perspectives on the current situation from the point of view of public reaction. These twitter datasets are open source and so can be gathered the largest quantities of tweets of the users. However, physicians and researchers also get various kinds of information that help them to get proper knowledge about this and explore innovative things to prevent this pandemic.

## 4. Conclusion

In this work, we proposed a clustered based classification and topics extraction model named TClustVID that can produce improved results of the classifiers compared to the traditional methods, and extracted significant topics from the high performing clusters of COVID-19 twitter datasets. This is almost the first study in which COVID-19-related twitter data has been investigated using this proposed model where different machine learning algorithms show the best results compared to more traditional approaches. In this work, TClustVID generates several clusters where one of them represent high classification accuracy that means it contains more significant topics that really represents the public opinions on twitter. In TClustVID, we used most widely used k-means clustering [14], when the contents of primary data are merged with clustered groups again and retokenizes this process. So, it makes the best appropriate results and helps machine learning classifier to understand different categories more clearly. In addition, this study not only identified the best classification model but also extracted significant topics that could be used for designing strategies to counter the pandemic. A great deal of information can be abstracted from very large numbers of tweets by the extraction of commonly occurring topics using LDA. These knowledge can be extracted from positive, neutral and negative tweets and identified high frequent information that are being transmitted and commented as the response to the pandemic situation. Such information can also use to gain insight into population activities, demands, opinions and responsibilities, and might be used to trace otherwise unidentified hot spots of COVID-19 infection by investigating topics and its categories and cross correlating this with medical data from other sources. There are some important limitations to note, such as these datasets do not contain more instances in upcoming months that relate to the COVID-19 tweets on twitter. Again, the interpretation of topics is a challenging task, hence some manual interpretation of topics may misinterpret in the topics modeling. In future work, more COVID-19 twitter data will be collected from different data repositories and investigated with these and other more advanced techniques currently being developed, which will enable more significant information extraction on COVID-19 topics.

## Data Availability

Experimental data has been gathered from IEEE Dataport

## Notes

### Competing Interest Statement

The authors have declared no competing interest.

### Funding Statement

No external funding was received

### Author Declarations

All relevant ethical guidelines have been followed

